# Assessing the Role of Red Blood Cell Distribution Width in Hospital Mortality Among Elderly and Non-Elderly COVID-19 Patients: A Prospective Study in a Brazilian University Hospital

**DOI:** 10.1101/2024.08.05.24311491

**Authors:** Helena Duani, Máderson Alvares de Souza Cabral, Carla Jorge Machado, Thalyta Nogueira Fonseca, Milena Soriano Marcolino, Vandack Alencar Nobre, Cecilia Gómez Ravetti, Paula Frizera Vassallo, Unaí Tupinambás

## Abstract

This study investigated the role of red blood cell distribution width (RDW) as a risk factor for hospital mortality in COVID-19 patients at a public hospital in Minas Gerais, Brazil. The study included 161 patients over 16 years old hospitalized between May and October 2020, with 39 (24.2%) deaths. Key mortality risk factors identified were age over 70 years (RR=2.78; p<0.001), male sex (RR=2.28; p=0.005), cardiovascular disease (RR=1.8; p=0.044), and abnormal chest X-ray upon admission (RR=3.07; p=0.022). Although high RDW at admission did not significantly predict mortality (31.1% vs 21.7%; RR=1.43; p=0.413), it was linked to higher mortality in patients aged 70 and over (66.7% vs 33.3%; RR=2; p=0.029). High RDW during hospitalization was a strong mortality factor for the entire cohort (41.1% vs 10.2%; RR=4.03; p<0.001) and at any time during the stay (39.7% vs 9.6%; RR=4.14; p<0.001). The Cox model analysis showed that age >70 years (HR=4.8; p<0.001), white race (HR=3.2; p=0.018), need for invasive ventilation (HR=3.8; p=0.001), and abnormal chest X-ray (HR=3.5; p=0.044) were significant risk factors, but RDW was not associated with mortality.

## Introduction

In December 2019, an outbreak of atypical cases of viral pneumonia, henceforth referred to as COVID-19, was reported in Wuhan, China. A few weeks later, a new beta-coronavirus – the severe acute respiratory syndrome coronavirus 2 (SARS-CoV-2) – was established as the etiologic agent of this disease. COVID-19 quickly spread throughout the globe and was classified as a pandemic by the World Health Organization in March 2020 [1].

The spectrum of clinical presentation of COVID-19 is broad, varying from asymptomatic infection to severe and critical disease. Patients with old age and comorbidities, such as heart, lung, or kidney disease; hypertension; diabetes; cancer; smoking; and obesity have a higher risk for severe disease and mortality. [2-7]

Many researchers are interested in identifying circulating biomarkers able to select patients with a higher risk of unfavorable clinical outcomes and death. In this context, some studies have shown that high D-dimer levels, lymphocytopenia [3,8], and some higher red blood cell distribution widths (RDW) are associated with poor outcomes [9]. Froy et al. concluded that patients with COVID-19 that presented with a high RDW upon hospital admission and an increasing RDW during hospitalization presented a higher risk of mortality [9].

Thus, the present study aimed to investigate, in a Brazilian sample of COVID-19 patients admitted to a public university hospital, if RDW measured upon admission or during hospitalization is associated with hospital mortality.

## Material and Methods

This study included all patients aged ≥ 16 years who had been admitted between May 1 and October 31, 2020 to the Hospital das Clínicas of the Universidade Federal de Minas Gerais (HC-UFMG), in Belo Horizonte, Brazil, with Severe Acute Respiratory Syndrome (SARS) or clinical symptoms whose evolution led to the suspicion of COVID-19 within the first three weeks of hospitalization. Patients hospitalized for other pathologies who became infected with COVID-19 during hospitalization were not included. Belo Horizonte, the capital city of the Southeast region of Brazil, has a population of over 2 million people. Among these residents, more than 80,000 are aged between 70 and 74 years old, highlighting a significant elderly demographic within the city (https://cidades.ibge.gov.br/brasil/mg/belo-horizonte/panorama, assessed in July 11, 2024.

The HC-UFMG is a university hospital with more than 400 beds and is a regional reference for the treatment of high complexity cases, including organ transplants and neoplasms. Since the onset of the COVID-19 pandemic, the hospital has been a part of the Brazilian Unified Health System (SUS) for the treatment of suspected cases in the metropolitan area of Belo Horizonte, Brazil.

Patients were followed up by the study team through a daily active search. Data were collected prospectively based on the hospital’s electronic medical records and radiological and laboratory examination information systems.

A confirmed case of COVID-19 was defined as patients with a positive real-time polymerase chain reaction (RT-PCR) for the detection of SARS-CoV-2 in tracheal aspirate or naso-oropharyngeal swab. Cases with clinical suspicion and radiological findings but without laboratory confirmation were not included.

A questionnaire was filled out during the study team’s follow-up to obtain the variables to be studied and compiled in a database. The race/color variable was obtained by self-declaration.

The duration of symptoms reported in the first medical record was considered. Comorbidities reported in medical records were recorded and categorized. Immunosuppression was defined as the presence of neoplasia, an immunosuppressive hematologic condition, primary or acquired immunodeficiency, or a relevant clinical condition whose treatment involves the use of immunosuppressants or high-dose of corticosteroids. The first chest X-ray or computed tomography (CT) scan performed during hospitalization was considered. X-rays were considered suggestive if bilateral diffuse interstitial infiltrate was found in an independent review by two physicians (in case of disagreement, a third physician reviewed the exam for a definitive opinion) and CT findings were taken from an official report prepared by a radiologist.

All laboratory tests were performed according to clinical indication, following the institution’s protocol for the treatment of suspected COVID-19 cases. The admission blood count was defined as the first blood count test performed within 24 hours of admission. For this test, the values of the RDW, total leukocytes, lymphocytes, neutrophils, mean corpuscular volume (MCV), and platelets were analyzed as continuous variables and categorized as low, normal, or high (in relation to standardized reference values in the service laboratory). The RDW was also categorized as normal (no changes during hospitalization) or high (value above the upper limit of normality at some point during hospitalization).

This research was conducted in accordance with the ethical principles established in the Declaration of Helsinki. The study protocol was reviewed and approved by the Institutional Review Board of UFMG, under the approval number 30437020.9.3001.5124. All participants were informed about the study’s objectives, procedures, potential risks, and benefits, and they signed a written informed consent form prior to participation.

### Statistical Analysis

After categorizing the variables, an analysis stratified by survival or death, was performed using mean and standard deviation for continuous variables, with normal distribution, and absolute numbers and percentages for categorical variables. The adopted significance level was 5% (p<0.05).

For data stratified into normal RDW and high RDW groups, the percentage of deaths within these groups was calculated and compared across two selected age categories (<70 years and ≥70 years). The rationale for using 70 years as the age criterion was to investigate potential differences in RDW-related outcomes across age groups. This threshold is commonly employed in clinical studies to distinguish between younger and older populations, considering the physiological and clinical changes that occur with aging. Additionally, 72 years is recognized as the global life expectancy [10]. Other studies on COVID-19 mortality and comorbidities have shown that individuals aged 70 years or older are more likely to have underlying health conditions and, therefore, a higher risk of mortality [11].

Subsequently, the Gross and Mantel-Haenszel-adjusted risk ratios were calculated to assess the association between RDW levels and mortality risk. The same analytical procedure was applied to data stratified based on RDW upon admission (normal RDW upon admission vs. high RDW upon admission). A significance level of 5% (p<0.05) was adopted for all statistical tests.

Next, for the data stratified into normal RDW and high RDW, the percentage of deaths in these groups was calculated, which was then compared within two selected age groups (<70 years and 70 years or more), after which the Gross and Mantel-Haenszel-adjusted risk ratio was calculated. The same procedure was also used for data stratified into normal RDW upon admission and high RDW upon admission. The adopted significance level was 5% (p<0.05).

A time-to-death analysis was performed for variables that proved to be significant (p<0.05) in the comparison between people who survived and died using the Cox proportional hazards model in univariate regression analysis. Those variables that revealed a significance level equal to or less than 20% (p<0.20) formed the basis of an initial multivariate model, and only those with a significance level equal to or greater than 5% remained in this model (p<0.05). Finally, Kaplan Meier survival curves for the variables in the final model were estimated to obtain a better graphic visualization of the differences between the categories in time until death.

## Results

Among the 169 patients with laboratory confirmations of COVID-19 who were hospitalized during the study period, 161 patients were included, of which 145 (90.1%) were directly admitted to respiratory isolation with SARS and 16 (9.9%) were admitted with nonspecific symptoms that led to the suspicion of COVID-19 after an average of 6.6 (1 to 18) days. Eight patients with detectable RT-PCR were not included because they were suspected cases of healthcare-associated COVID-19 infection, it means that the patient acquired the virus inside the hospital, we not exclude another bacterial or fungal healthcare infections. The main characteristics presented by the patients included in the study are shown in Table 1. Of the 161 patients included, 39 (24.2%) died during hospitalization, 32 of them within the first 30 days, resulting in a 30-day mortality of 19.9%.

**Table 1:**
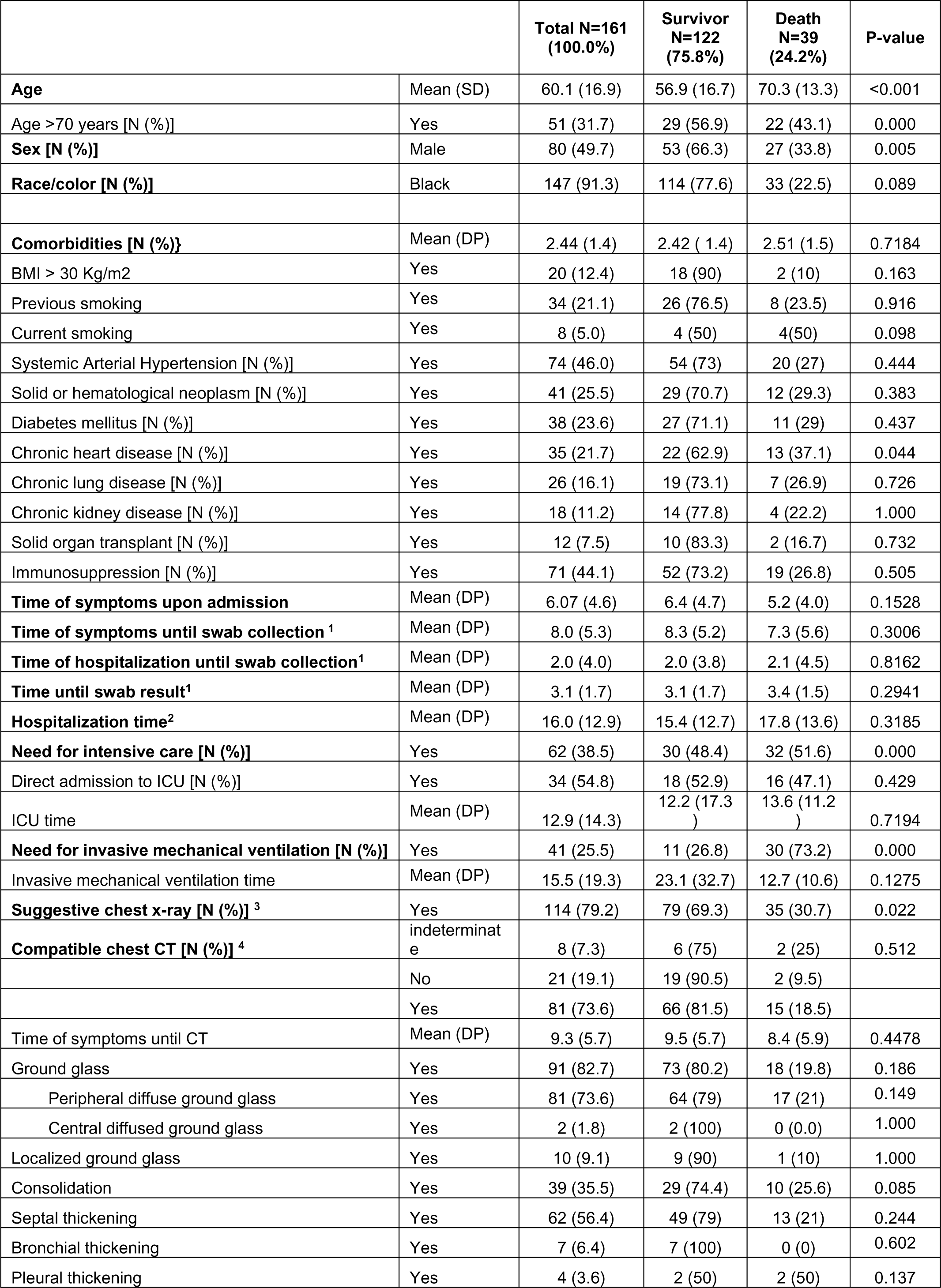
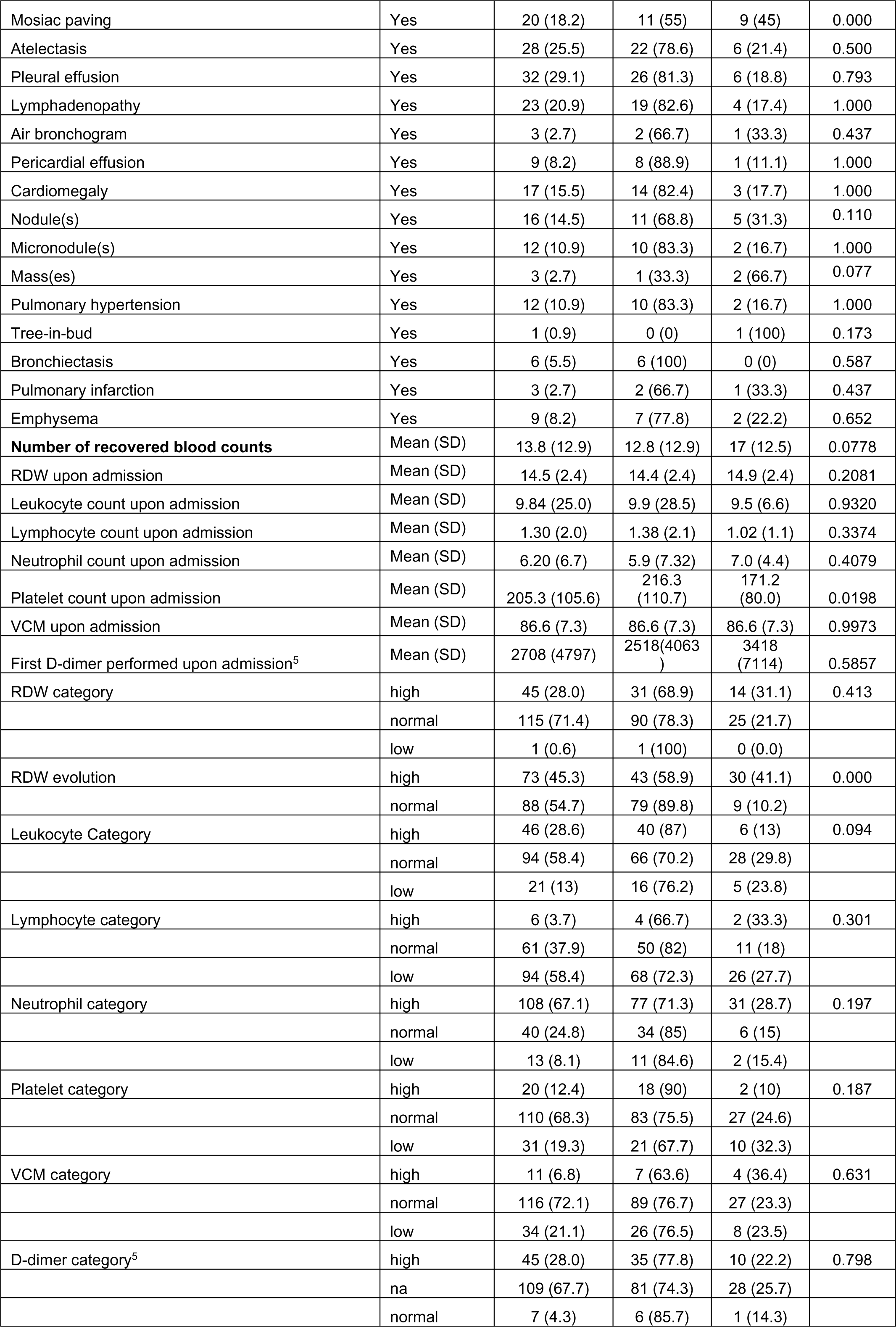
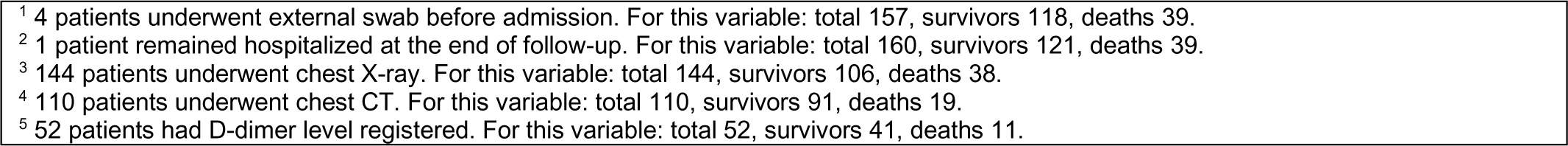
Main baseline characteristics, comorbidities, laboratory and radiological findings of COVID-19 patients, survivor and non-survivors, in Belo Horizonte, Brazil, 2020.

|  |  | Total N=161<br>(100.0%) | Survivor<br>N=122<br>(75.8%) | Death<br>N=39<br>(24.2%) | P-value |
| --- | --- | --- | --- | --- | --- |
| **Age** | Mean (SD) | 60.1 (16.9) | 56.9 (16.7) | 70.3 (13.3) | <0.001 |
| Age >70 years [N (%)] | Yes | 51 (31.7) | 29 (56.9) | 22 (43.1) | 0.000 |
| **Sex [N (%)]** | Male | 80 (49.7) | 53 (66.3) | 27 (33.8) | 0.005 |
| **Race/color [N (%)]** | Black | 147 (91.3) | 114 (77.6) | 33 (22.5) | 0.089 |
| **Comorbidities [N (%)]** | Mean (DP) | 2.44 (1.4) | 2.42 (1.4) | 2.51 (1.5) | 0.7184 |
| BMI > 30 Kg/m<sup>2</sup> | Yes | 20 (12.4) | 18 (90) | 2 (10) | 0.163 |
| Previous smoking | Yes | 34 (21.1) | 26 (76.5) | 8 (23.5) | 0.916 |
| Current smoking | Yes | 8 (5.0) | 4 (50) | 4 (50) | 0.098 |
| Systemic Arterial Hypertension [N (%)] | Yes | 74 (46.0) | 54 (73) | 20 (27) | 0.444 |
| Solid or hematological neoplasm [N (%)] | Yes | 41 (25.5) | 29 (70.7) | 12 (29.3) | 0.383 |
| Diabetes mellitus [N (%)] | Yes | 38 (23.6) | 27 (71.1) | 11 (29) | 0.437 |
| Chronic heart disease [N (%)] | Yes | 35 (21.7) | 22 (62.9) | 13 (37.1) | 0.044 |
| Chronic lung disease [N (%)] | Yes | 26 (16.1) | 19 (73.1) | 7 (26.9) | 0.726 |
| Chronic kidney disease [N (%)] | Yes | 18 (11.2) | 14 (77.8) | 4 (22.2) | 1.000 |
| Solid organ transplant [N (%)] | Yes | 12 (7.5) | 10 (83.3) | 2 (16.7) | 0.732 |
| Immunosuppression [N (%)] | Yes | 71 (44.1) | 52 (73.2) | 19 (26.8) | 0.505 |
| **Time of symptoms upon admission** | Mean (DP) | 6.07 (4.6) | 6.4 (4.7) | 5.2 (4.0) | 0.1528 |
| **Time of symptoms until swab collection <sup>1</sup>** | Mean (DP) | 8.0 (5.3) | 8.3 (5.2) | 7.3 (5.6) | 0.3006 |
| **Time of hospitalization until swab collection<sup>1</sup>** | Mean (DP) | 2.0 (4.0) | 2.0 (3.8) | 2.1 (4.5) | 0.8162 |
| **Time until swab result<sup>1</sup>** | Mean (DP) | 3.1 (1.7) | 3.1 (1.7) | 3.4 (1.5) | 0.2941 |
| **Hospitalization time<sup>2</sup>** | Mean (DP) | 16.0 (12.9) | 15.4 (12.7) | 17.8 (13.6) | 0.3185 |
| **Need for intensive care [N (%)]** | Yes | 62 (38.5) | 30 (48.4) | 32 (51.6) | 0.000 |
| Direct admission to ICU [N (%)] | Yes | 34 (54.8) | 18 (52.9) | 16 (47.1) | 0.429 |
| ICU time | Mean (DP) | 12.9 (14.3) | 12.2 (17.3) | 13.6 (11.2) | 0.7194 |
| **Need for invasive mechanical ventilation [N (%)]** | Yes | 41 (25.5) | 11 (26.8) | 30 (73.2) | 0.000 |
| Invasive mechanical ventilation time | Mean (DP) | 15.5 (19.3) | 23.1 (32.7) | 12.7 (10.6) | 0.1275 |
| **Suggestive chest x-ray [N (%)] <sup>3</sup>** | Yes | 114 (79.2) | 79 (69.3) | 35 (30.7) | 0.022 |
| **Compatible chest CT [N (%)] <sup>4</sup>** | indeterminate | 8 (7.3) | 6 (75) | 2 (25) | 0.512 |
|  | No | 21 (19.1) | 19 (90.5) | 2 (9.5) |  |
|  | Yes | 81 (73.6) | 66 (81.5) | 15 (18.5) |  |
| Time of symptoms until CT | Mean (DP) | 9.3 (5.7) | 9.5 (5.7) | 8.4 (5.9) | 0.4478 |
| Ground glass | Yes | 91 (82.7) | 73 (80.2) | 18 (19.8) | 0.186 |
| Peripheral diffuse ground glass | Yes | 81 (73.6) | 64 (79) | 17 (21) | 0.149 |
| Central diffused ground glass | Yes | 2 (1.8) | 2 (100) | 0 (0.0) | 1.000 |
| Localized ground glass | Yes | 10 (9.1) | 9 (90) | 1 (10) | 1.000 |
| Consolidation | Yes | 39 (35.5) | 29 (74.4) | 10 (25.6) | 0.085 |
| Septal thickening | Yes | 62 (56.4) | 49 (79) | 13 (21) | 0.244 |
| Bronchial thickening | Yes | 7 (6.4) | 7 (100) | 0 (0) | 0.602 |
| Pleural thickening | Yes | 4 (3.6) | 2 (50) | 2 (50) | 0.137 |
| Mosaic paving | Yes | 20 (18.2) | 11 (55) | 9 (45) | 0.000 |
| Atelectasis | Yes | 28 (25.5) | 22 (78.6) | 6 (21.4) | 0.500 |
| Pleural effusion | Yes | 32 (29.1) | 26 (81.3) | 6 (18.8) | 0.793 |
| Lymphadenopathy | Yes | 23 (20.9) | 19 (82.6) | 4 (17.4) | 1.000 |
| Air bronchogram | Yes | 3 (2.7) | 2 (66.7) | 1 (33.3) | 0.437 |
| Pericardial effusion | Yes | 9 (8.2) | 8 (88.9) | 1 (11.1) | 1.000 |
| Cardiomegaly | Yes | 17 (15.5) | 14 (82.4) | 3 (17.7) | 1.000 |
| Nodule(s) | Yes | 16 (14.5) | 11 (68.8) | 5 (31.3) | 0.110 |
| Micronodule(s) | Yes | 12 (10.9) | 10 (83.3) | 2 (16.7) | 1.000 |
| Mass(es) | Yes | 3 (2.7) | 1 (33.3) | 2 (66.7) | 0.077 |
| Pulmonary hypertension | Yes | 12 (10.9) | 10 (83.3) | 2 (16.7) | 1.000 |
| Tree-in-bud | Yes | 1 (0.9) | 0 (0) | 1 (100) | 0.173 |
| Bronchiectasis | Yes | 6 (5.5) | 6 (100) | 0 (0) | 0.587 |
| Pulmonary infarction | Yes | 3 (2.7) | 2 (66.7) | 1 (33.3) | 0.437 |
| Emphysema | Yes | 9 (8.2) | 7 (77.8) | 2 (22.2) | 0.652 |
| **Number of recovered blood counts** | Mean (SD) | 13.8 (12.9) | 12.8 (12.9) | 17 (12.5) | 0.0778 |
| RDW upon admission | Mean (SD) | 14.5 (2.4) | 14.4 (2.4) | 14.9 (2.4) | 0.2081 |
| Leukocyte count upon admission | Mean (SD) | 9.84 (25.0) | 9.9 (28.5) | 9.5 (6.6) | 0.9320 |
| Lymphocyte count upon admission | Mean (SD) | 1.30 (2.0) | 1.38 (2.1) | 1.02 (1.1) | 0.3374 |
| Neutrophil count upon admission | Mean (SD) | 6.20 (6.7) | 5.9 (7.32) | 7.0 (4.4) | 0.4079 |
| Platelet count upon admission | Mean (SD) | 205.3 (105.6) | 216.3 (110.7) | 171.2 (80.0) | 0.0198 |
| VCM upon admission | Mean (SD) | 86.6 (7.3) | 86.6 (7.3) | 86.6 (7.3) | 0.9973 |
| First D-dimer performed upon admission<sup>5</sup> | Mean (SD) | 2708 (4797) | 2518(4063 ) | 3418 (7114) | 0.5857 |
| RDW category | high | 45 (28.0) | 31 (68.9) | 14 (31.1) | 0.413 |
|  | normal | 115 (71.4) | 90 (78.3) | 25 (21.7) |  |
|  | low | 1 (0.6) | 1 (100) | 0 (0.0) |  |
| RDW evolution | high | 73 (45.3) | 43 (58.9) | 30 (41.1) | 0.000 |
|  | normal | 88 (54.7) | 79 (89.8) | 9 (10.2) |  |
| Leukocyte Category | high | 46 (28.6) | 40 (87) | 6 (13) | 0.094 |
|  | normal | 94 (58.4) | 66 (70.2) | 28 (29.8) |  |
|  | low | 21 (13) | 16 (76.2) | 5 (23.8) |  |
| Lymphocyte category | high | 6 (3.7) | 4 (66.7) | 2 (33.3) | 0.301 |
|  | normal | 61 (37.9) | 50 (82) | 11 (18) |  |
|  | low | 94 (58.4) | 68 (72.3) | 26 (27.7) |  |
| Neutrophil category | high | 108 (67.1) | 77 (71.3) | 31 (28.7) | 0.197 |
|  | normal | 40 (24.8) | 34 (85) | 6 (15) |  |
|  | low | 13 (8.1) | 11 (84.6) | 2 (15.4) |  |
| Platelet category | high | 20 (12.4) | 18 (90) | 2 (10) | 0.187 |
|  | normal | 110 (68.3) | 83 (75.5) | 27 (24.6) |  |
|  | low | 31 (19.3) | 21 (67.7) | 10 (32.3) |  |
| VCM category | high | 11 (6.8) | 7 (63.6) | 4 (36.4) | 0.631 |
|  | normal | 116 (72.1) | 89 (76.7) | 27 (23.3) |  |
|  | low | 34 (21.1) | 26 (76.5) | 8 (23.5) |  |
| D-dimer category<sup>5</sup> | high | 45 (28.0) | 35 (77.8) | 10 (22.2) | 0.798 |
|  | na | 109 (67.7) | 81 (74.3) | 28 (25.7) |  |
|  | normal | 7 (4.3) | 6 (85.7) | 1 (14.3) |  |

<sup>1</sup> 4 patients underwent external swab before admission. For this variable: total 157, survivors 118, deaths 39. <sup>2</sup> 1 patient remained hospitalized at the end of follow-up. For this variable: total 160, survivors 121, deaths 39. <sup>3</sup> 144 patients underwent chest X-ray. For this variable: total 144, survivors 106, deaths 38. <sup>4</sup> 110 patients underwent chest CT. For this variable: total 110, survivors 91, deaths 19. <sup>5</sup> 52 patients had D-dimer level registered. For this variable: total 52, survivors 41, deaths 11.

Eighty patients (49.7%) were male and 147 (91.3%) were self-declared black (black or brown). Mortality was 14.8% among women and 33.8% among men, this difference being statistically significant (RR=2.28; p=0.005). There was no difference in mortality rates between whites and blacks (42.7% vs 22.5%; RR=1.9; p=0.089).

The mean age was 60.1 years (17 to 93, standard deviation - SD 16.9), with a median of 63 years. The mean age among survivors was 56.9 (16.7) years, while it was 70.3 (13.3) years among non-survivors, with a statistically significant difference (p<0.001). In addition, patients over 70 years of age had a higher mortality rate (43.1% vs 15.5%; RR=2.78; p<0.001).

The mean time from symptoms to hospital admission was 6.1 days (standard deviation - SD - 4.6), with a median of 5 days. Patients had an average of 2.44 comorbidities (SD 1.4), the most common being systemic arterial hypertension (74; 46%) and solid or hematologic malignancy (41; 25.5%). Seventy-one patients (44.1%) were considered to have immunosuppression. Four patients were living with HIV. Only eight (5%) patients had no previous comorbidity, with 38 (23.6%) having one, 45 (27.9%) two, and 70 (43.5%) at least three comorbidities.

Among the most commonly reported comorbidities, the presence of chronic heart disease was a risk factor for mortality (37.1% vs 20.6%; RR=1.8; p=0.044). The presence of other comorbidities was not associated with mortality.

In this cohort, 62 patients (38.5%) developed a critical form of the disease and required admission to intensive care, with 34 of them (54.8%) being admitted directly to the Intensive Care Unit (ICU). The mean length of stay in ICU was 12.9 (14.3) days. Forty-one patients (25.5%) required invasive mechanical ventilation (MV), with a mean length of stay of 15.5 (19.3) days.

As expected, the mortality rate was higher among patients who required intensive care (51.6% vs 7.1%; RR= 7.26; p<0.001) and those who required mechanical ventilation (73.2% vs 7.3%; RR=10.03; p< 0.001). However, there was no difference in mortality rates between patients who were admitted directly to intensive care and those who were admitted to the ward and later transferred to intensive care (47.1% vs 57.1%; RR=0.82; p=0.429). Furthermore, among those who required such interventions, there was no difference in the length of stay in intensive care (mean 13.6, SD 11.2 vs mean 12.2, SD 17.3; p=0.7194) or in the time of MV (mean 12.7, SD 10.6 vs mean 23.1, SD 32.7; p=0.1275) between deaths and survivors.

### Imaging exams

One hundred and forty-four (89.4%) patients underwent at least one chest X-ray during hospitalization, the first being analyzed in cases in which it was performed more than once. COVID-19 was considered suggestive in 114 (79.2%) of these patients based on an interpretation by two clinicians.

Computed tomography of the chest was performed in 110 (68.3%) of the patients, on average 9.3 days after the onset of symptoms (SD 5.7), with the results being compatible with COVID-19 in 81 cases (73.6%) in an official report prepared by a radiologist. The most common findings were: ground glass opacities in 91 cases (82.7%), which was diffuse with peripheral predominance in 81 cases (73.6%; 2 of these patients also had ground glass opacities of bilateral central distribution) and localized in 10 cases (9.1%); septal thickening in 62 cases (56.4%); consolidation in 39 cases (35.5%); pleural effusion in 32 cases (29.1%); atelectasis in 28 cases (25.5%); lymphadenopathy in 23 cases (20.9%); and mosaic paving in 20 cases (18.2%).

A chest X-ray considered suggestive of COVID-19 was associated with a higher mortality rate (30.7% vs 10%; RR=3.07; p=0.022). However, a chest CT that was reported as compatible with COVID-19 did not represent an additional risk of mortality (18.5% vs 9.5%; RR=1.95; p=0.512). Among the findings described in the CT scan reports, the presence of a mosaic paving pattern was found to be a risk factor for higher mortality (45% vs 11.1%; RR=4.05; p<0.001). No difference was found in the time taken to perform a chest CT between survivors and deaths (mean 9.5, SD 5.7 vs mean 8.4, SD 5.9; p=0.4478).

### Laboratory exams

Fifty-two patients underwent at least one D-dimer measurement at some point during hospitalization, with mean values of 2,708 (SD 4,797), which was categorized as normal in 7 (13.5%) and high in 45 (86.5%) patients.

No difference was found in the D-dimer value between survivors and non-survivors (mean 2518, SD 4,063 vs mean 3418, SD 7,114; p=0.5857) and having a high D-dimer was not a risk factor for mortality (22.2% vs 14.3%; RR=1.55; p=0.798).

All 161 patients underwent at least one blood count test during hospitalization. A total of 2,227 blood counts were taken, with a mean of 13.8 per patient (standard deviation 12.9), ranging from 1 to 84. The RDW upon admission was normal in 115 (71.4%), high in 45 (28%), and low in 1 (0.6%). The mean RDW values upon admission were 14.5 (SD 2.4), a value that is within the normal range.

Among patients with normal RDW upon admission, 82 (71.3%) remained normal and 33 (28.7%) rose to high levels over time. Among those with high RDW upon admission, 40 (88.9%) remained high and 5 (11.1%) returned to normal. The only patient with low RDW upon admission returned to normal. When categorized, 88 (54.7%) had normal RDW and 73 (45.3%) had high RDW during the whole hospitalization time. No difference was found in the mean RDW value upon admission between survivors and non-survivors (14.4 [2.4] between survivors’ vs 14.9 [2.4] between non-survivors; p=0.2081).

The risk for mortality in the group with high RDW upon admission was higher than in the group with normal RDW, but without statistical significance (31.1% vs 21.7%; RR=1.43; p=0.413). However, in the subgroup of patients ≥70 years old, a statistical significance was observed in the mortality (66.7% for high RDW vs 33.3% for normal RDW; RR=2; p=0.029), Table 2.

**Table 2:** Mortality risk according to age-stratified RDW category (normal or high) of COVID-19 patients, in Belo Horizonte, Brazil, 2020.

| Age group | Normal RDW during hospitalization |  | High RDW during hospitalization |  | P-value | Risk ratio (95% CI) |
| --- | --- | --- | --- | --- | --- | --- |
|  | N | Mortality [n (%)] | N | Mortality [n (%)] |  |  |
| <70 | 59 | 4 (6.8) | 51 | 13 (25.5) | 0.007 | 3.76 (1.31;10.8) |
| 70+ | 24 | 4 (16.7) | 27 | 18 (66.7) | <0.001 | 4.00 (1.58;10.2) |
| **Total cohort** | 83 | 8 (9.6) | 78 | 31 (39.7) | <0.001 |  |
| gross |  |  |  |  |  | 4.12 (2.02; 8.41) |
| Mantel-Haenszel Method |  |  |  |  |  | 3.89 (1.93; 7.83) |
| Age group | Normal RDW at admission |  | High RDW at admission |  | P- value | Risk ratio (95% CI) |
|  | N | Mortality [n (%)] | N | Mortality [n (%)] |  |  |
| <70 | 80 | 13 (16.3) | 30 | 4 (13.3) | >0.999 | 0.82 (0.29;2.31) |
| 70+ | 36 | 12 (33.3) | 15 | 10 (66.7) | 0.029 | 2.00 (1.07;3.74) |
| **Total cohort** | 116 | 25 (21.6) | 45 | 14 (31.1) | 0.204 |  |
| gross |  |  |  |  |  | 1.44 (0.82; 2.55) |
| Mantel-Haenszel Method |  |  |  |  |  | 1.41 (0.83; 2.39) |

Furthermore, when subsequent laboratory findings of progression (RDW evolution, Table 1) is categorized, there is a higher risk of mortality among those with high RDW after admission (41.1% vs 10.2%; RR=4.03; p<0.001). Moreover, when considering the RDW that changes at any time during hospitalization against such a normal parameter in all of the blood count results, a significant risk of mortality tends to be observed for the entire cohort (39.7% vs 9.6%; RR=4.14; p<0.001).

No significant difference was found in mortality between the different categories upon admission in relation to other parameters analyzed in the blood count, except for the absolute platelet counts between survivors and non-survivors (mean 216.3x10^3^, SD 110.7 vs mean 171.2x10^3^, SD 80; p=0.0198).

### Survival analysis

In a univariate analysis using the Cox Model (Table 3), the risk factors for lower survival were: male sex, HR 2.3 (1.1;4.5), p=0.018; age >70 years, HR 3.3 (1.7;6.5), p<0.001; white race, HR 2.5 (1.0;6.1), p=0.039; need for invasive mechanical ventilation, HR 4.9 (2.2;10.6), p<0.001; and need for intensive care admission, HR 3.7 (1.6;8.6), p=0.003.

**Table 3.**
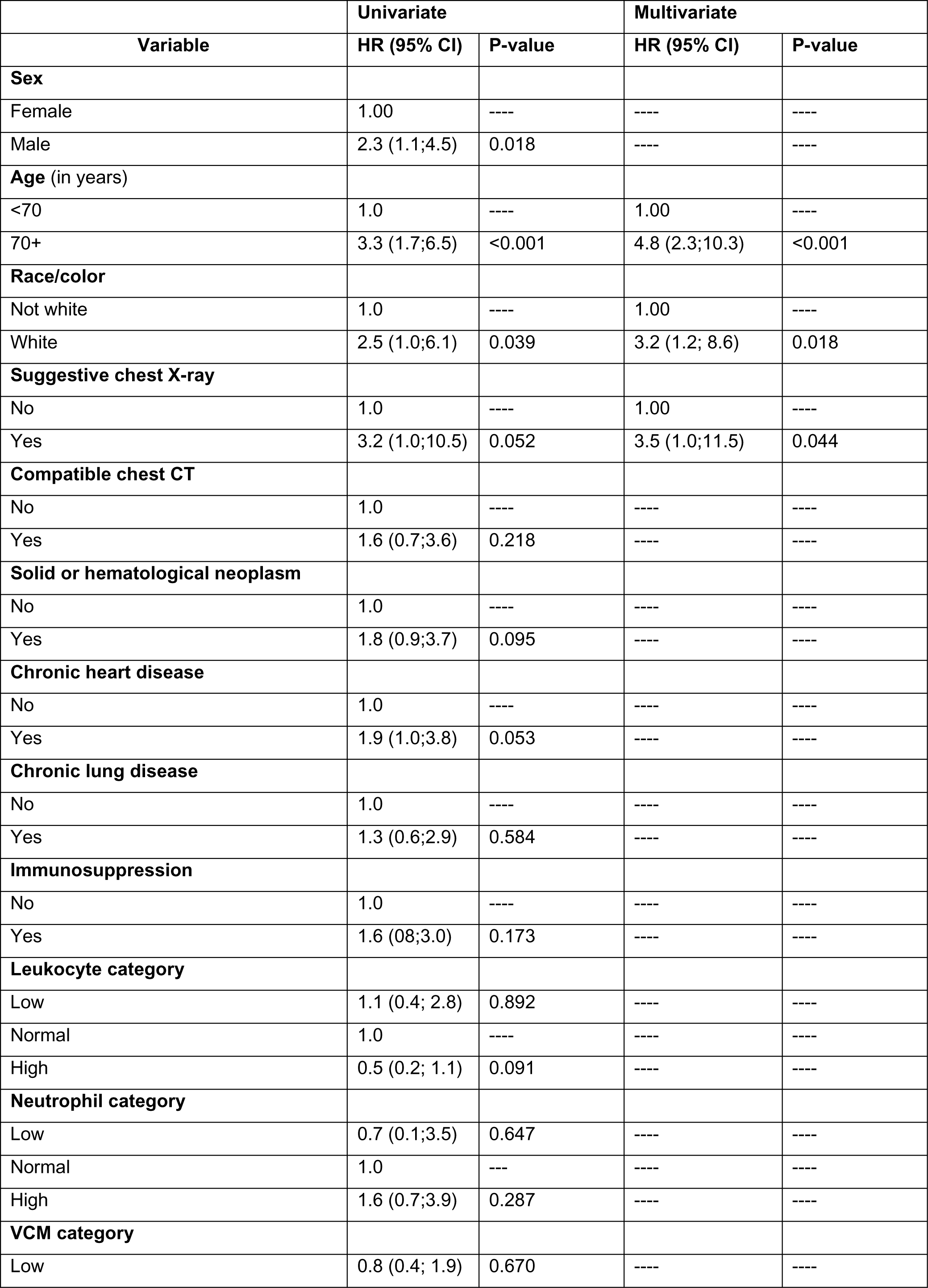
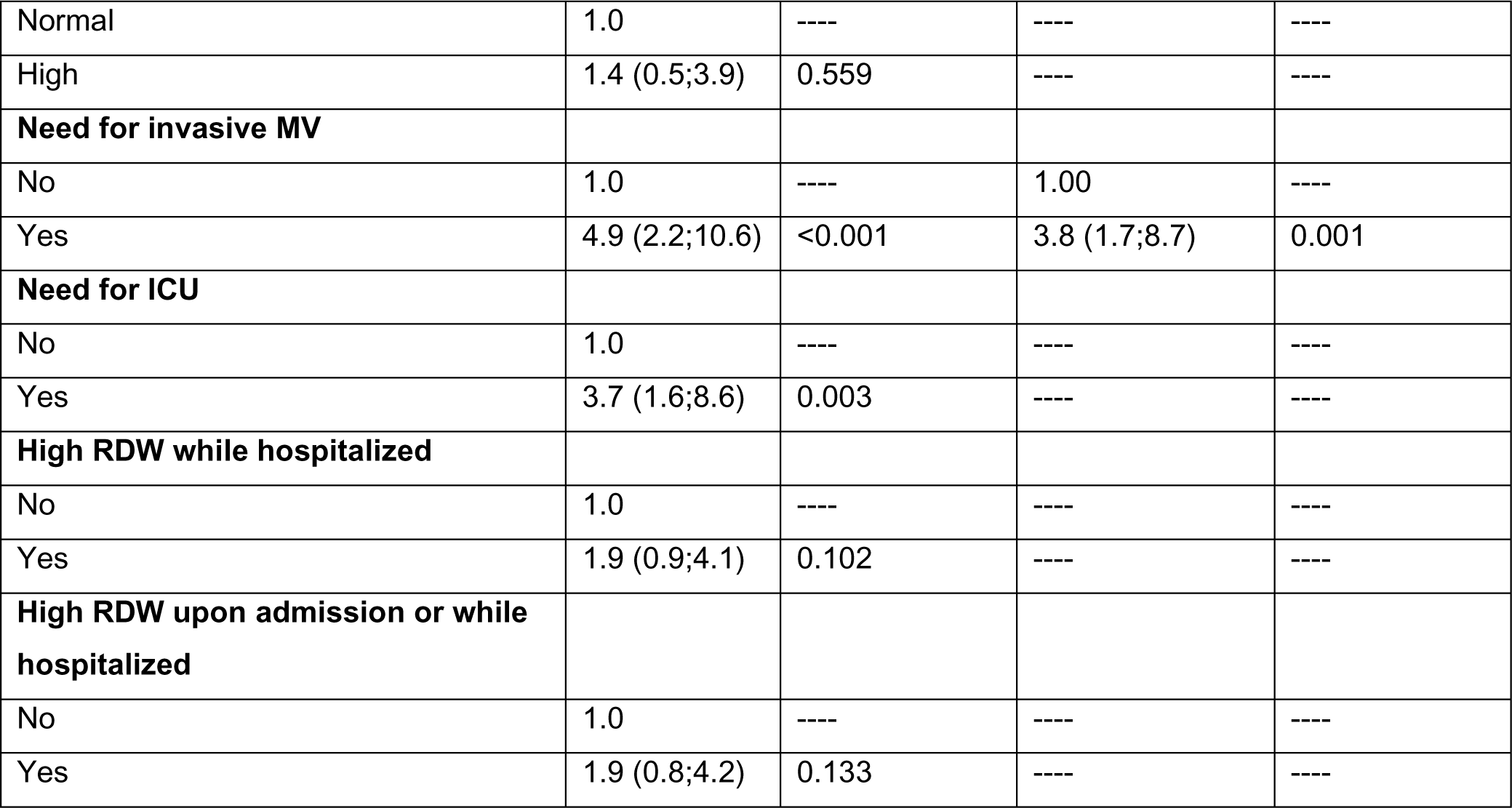
Cox Model results and survival analysis of baseline characteristics, comorbidities, laboratory and radiological findings of COVID-19 patients, survivor and non-survivors, in Belo Horizonte, Brazil, 2020.

| Variable | Univariate |  | Multivariate |  |
| --- | --- | --- | --- | --- |
|  | HR (95% CI) | P-value | HR (95% CI) | P-value |
| **Sex** |  |  |  |  |
| Female | 1.00 | ---- | ---- | ---- |
| Male | 2.3 (1.1;4.5) | 0.018 | ---- | ---- |
| **Age (in years)** |  |  |  |  |
| <70 | 1.0 | ---- | 1.00 | ---- |
| 70+ | 3.3 (1.7;6.5) | <0.001 | 4.8 (2.3;10.3) | <0.001 |
| **Race/color** |  |  |  |  |
| Not white | 1.0 | ---- | 1.00 | ---- |
| White | 2.5 (1.0;6.1) | 0.039 | 3.2 (1.2; 8.6) | 0.018 |
| **Suggestive chest X-ray** |  |  |  |  |
| No | 1.0 | ---- | 1.00 | ---- |
| Yes | 3.2 (1.0;10.5) | 0.052 | 3.5 (1.0;11.5) | 0.044 |
| **Compatible chest CT** |  |  |  |  |
| No | 1.0 | ---- | ---- | ---- |
| Yes | 1.6 (0.7;3.6) | 0.218 | ---- | ---- |
| **Solid or hematological neoplasm** |  |  |  |  |
| No | 1.0 | ---- | ---- | ---- |
| Yes | 1.8 (0.9;3.7) | 0.095 | ---- | ---- |
| **Chronic heart disease** |  |  |  |  |
| No | 1.0 | ---- | ---- | ---- |
| Yes | 1.9 (1.0;3.8) | 0.053 | ---- | ---- |
| **Chronic lung disease** |  |  |  |  |
| No | 1.0 | ---- | ---- | ---- |
| Yes | 1.3 (0.6;2.9) | 0.584 | ---- | ---- |
| **Immunosuppression** |  |  |  |  |
| No | 1.0 | ---- | ---- | ---- |
| Yes | 1.6 (0.8;3.0) | 0.173 | ---- | ---- |
| **Leukocyte category** |  |  |  |  |
| Low | 1.1 (0.4; 2.8) | 0.892 | ---- | ---- |
| Normal | 1.0 | ---- | ---- | ---- |
| High | 0.5 (0.2; 1.1) | 0.091 | ---- | ---- |
| **Neutrophil category** |  |  |  |  |
| Low | 0.7 (0.1;3.5) | 0.647 | ---- | ---- |
| Normal | 1.0 | --- | ---- | ---- |
| High | 1.6 (0.7;3.9) | 0.287 | ---- | ---- |
| **VCM category** |  |  |  |  |
| Low | 0.8 (0.4; 1.9) | 0.670 | ---- | ---- |
| Normal | 1.0 | ---- | ---- | ---- |
| High | 1.4 (0.5;3.9) | 0.559 | ---- | ---- |
| **Need for invasive MV** |  |  |  |  |
| No | 1.0 | ---- | 1.00 | ---- |
| Yes | 4.9 (2.2;10.6) | <0.001 | 3.8 (1.7;8.7) | 0.001 |
| **Need for ICU** |  |  |  |  |
| No | 1.0 | ---- | ---- | ---- |
| Yes | 3.7 (1.6;8.6) | 0.003 | ---- | ---- |
| **High RDW while hospitalized** |  |  |  |  |
| No | 1.0 | ---- | ---- | ---- |
| Yes | 1.9 (0.9;4.1) | 0.102 | ---- | ---- |
| **High RDW upon admission or while hospitalized** |  |  |  |  |
| No | 1.0 | ---- | ---- | ---- |
| Yes | 1.9 (0.8;4.2) | 0.133 | ---- | ---- |

We included the variables that were significant in the univariate analysis in our multivariate model. Although RDW was not included because it was significant only for a subset of the population (patients over 70 years), we aimed to develop a generalized model suitable for the diverse and complex patient population in our cohort. The multivariate analysis was adjusted for variables that provided the best model fit, enhancing its robustness despite the complexity of the cases studied. When performing a multivariate analysis, the following continue to be risk factors for lower survival: age >70 years, HR 4.8 (95% CI: 2.3;10.3), p<0.001; white race, HR 3.2 (95% CI: 1.2; 8.61), p=0.018; and need for invasive MV, HR 3.8 (1.7;8.7), p=0.001. The presence of a chest X-ray suggestive of COVID-19, HR 3.5 (95% CI: 1.0;11.5), p=0.044, also appears to be a risk factor in this analysis. Figure 1 shows the Kaplan-Meier curves for the variables that presented a lower risk of survival in the multivariate analysis.

**Figure 1:**
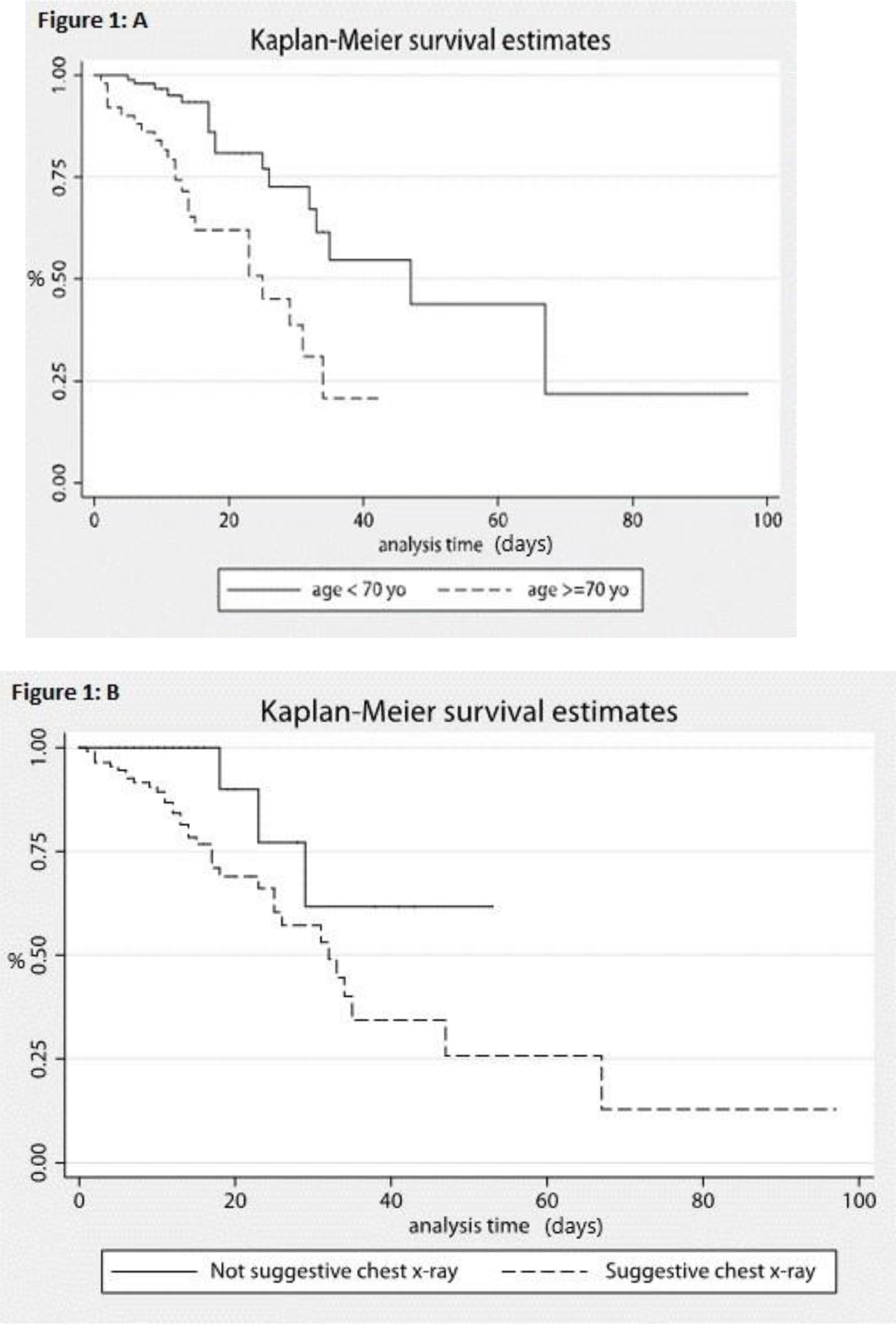
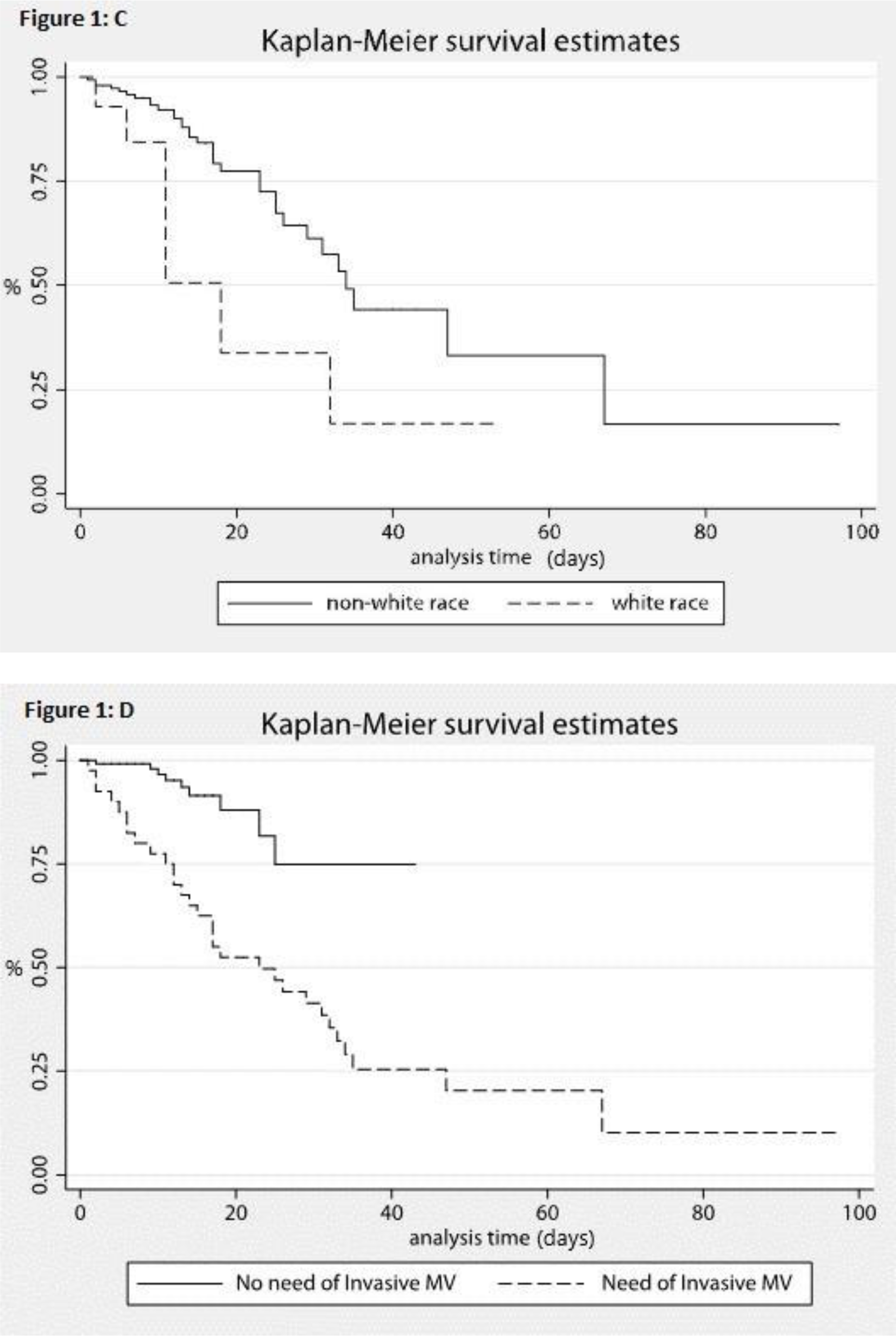
Kaplan-Meier curves for survival estimates of COVID-19 patients in a public university hospital in Belo Horizonte, Brazil, 2020. A Kaplan-Meier survival estimates by age: <70 and ≥70 years. B. Kaplan-Meier survival estimates by not suggestive chest x-ray and suggestive. C.Kaplan-Meier survival estimates by non-white race and white race. D. Kaplan-Meier survival estimates by non-need of invasive MV (mechanical ventilation) and need of MV.

## Discussion

This work studies the mortality rates of patients with confirmed COVID-19 who were admitted to a university hospital, a reference for the treatment of highly complex conditions. The objective was to find the factors that are unique to this population with different comorbidities, as well as the factors involved in greater severity and mortality from the disease.

The total mortality rate was 24.2% during hospitalization, with a 30-day mortality rate of 19.9%. Initial studies showed a mortality rate of 4.3% to 15% among patients hospitalized for SARS, [1,8], with more recent studies showing that it reaches 14% to 28% [3,12].

The data show a high median age (63 years) among patients hospitalized with SARS and that those in the older age group have a higher mortality rate (43.1% in those aged 70 years or over vs 15.5% in the others; RR=2.78; p<0.001). Initial cohorts studied in China found median ages of 49 to 56 years among confirmed and hospitalized cases of COVID-19, [1,8] although higher medians were also found in later studies, such as a cohort study in New York that found a median age of 63 years in patients with SARS [12]. Other studies have also shown higher mortality rates in older age strata [2,12,13]. When stratified by the same age groups, data from a multicenter study in New York found similar mortality rates (21% in-hospital mortality, with 42.83% aged 70 years or over vs 9.97% in other adults) [13]. A Brazilian multicenter study, including patients from less complex hospitals, found a mortality rate of 35.88% in patients aged 65 years or older vs 12.88% in the others [14].

In this study’s cohort, individuals had an average of 2.44 comorbidities before admission. This number is similar to that found in a subgroup of 355 patients in a report from Italy who died from COVID-19, where the average previous comorbidity was 2.7, [13] which reflects the previous clinical complexity of the patients included in this study. In particular, 95% of the individuals included in the present study had at least one previous comorbidity, with 23.6% having one, 27.9% having two, and 43.5% having at least 3 comorbidities. The Italian study showed a prevalence of comorbidity of 25.1%, 25.6%, and 48.5%, respectively, in patients who died from COVID-19 [13]. This reflects a greater previous clinical severity of patients treated in a high complexity hospital, which proved to be in the spectrum of the highest prevalence of comorbidities found in other studies. While there is a high prevalence of classic non-communicable comorbidities in other cohorts, such as arterial hypertension, obesity, and diabetes mellitus, [3,13,14] there was a high prevalence of more complex conditions in this study, such as neoplasms, heart disease, and immunosuppression, which are considered important factors for greater severity and mortality [5,13].

Males corresponded to 49.7% of the cases, which is a risk factor for mortality (33.8% vs 14.8%; RR=2.28; p=0.005). Males also account for a disproportionately high number of critical cases and deaths in diverse cohorts worldwide [13,15-17]. A Danish study showed a mortality rate among confirmed COVID-19 cases of 11.2% in males and 7.4% in females, with 47.1% of the individuals included being male [16]. A meta-analysis using data from 90 studies from 46 different countries totaling 3,111,714 individuals with a similar proportion of the sexes identified the male sex as a risk factor for 2.84 times more intensive care admissions and 1.39 times greater mortality [17].

The black race (self-declared black or brown individuals) represented more than 90% of this cohort, in contrast with the proportions found in the Brazilian population, where in the 2019 National Household Sample Survey (in Portuguese: Pesquisa Nacional por Amostra de Domicílios - PNAD), 42.7% of Brazilians declared themselves as white, 46.8% as brown, 9.4% as black, and 1.1% as yellow or indigenous [18]. This percentage may reflect disparities in the social determinants of health in this population segment, as was found in other studies [19, 20], in addition to highlighting social inequality in the Brazilian population. There is also a tendency toward a higher mortality rate among whites (although without statistical significance), with this group having a lower survival in the Cox model; however, the small percentage of self-declared white patients limits the extrapolation of these findings to the general population. Analyses controlled for comorbidities and socioeconomic factors found no association between race and in-hospital COVID-19 outcomes [21,22].

A median of 5 days of symptoms at the time of hospitalization was slightly lower than that found in other studies. Data suggest disease progression with mild symptoms in the first week [23]. One study showed a median of 5 days for the onset of dyspnea and 7 days for hospital admission [8]. Another study found a median of 8 days for the onset of dyspnea [1].

This study’s data also draws attention to one of the difficulties in the management of respiratory syndromes: the etiological diagnosis. Despite the collection being done shortly after admission, the test results of swab exams take an average of three days to be released. One study showed a mean time of 15.4 hours for RT-PCR results [12]. The delay in making the etiological diagnosis of pulmonary conditions can lead to the unnecessary use of antimicrobials and their consequences, as well as the need to use other more invasive forms of workup to clarify the diagnosis [24].

The need for intensive care in 38% of hospitalized patients in this cohort was compatible with the end of greater severity reported in other studies, where it varied between 14% and 48%, but with a similar mortality rate [3,12,13,14]. The need for MV of 25.5% was also similar to findings described in other studies, varying between 12% and 33% [3,12,14], but this time with a higher mortality rate among those requiring MV, which was 73% in our study compared to 60% in others [3]. The severity of the underlying health conditions in this cohort may have contributed to its occurrence since the patients were being treated for serious diseases in a high-complexity hospital. Despite not being statistically significant, the tendency for those who survived to spend a longer amount of time on mechanical ventilation is noteworthy, suggesting possible earlier death.

Having a chest X-ray taken – a simple, quick, and inexpensive procedure that can be easily interpreted by a radiologist – was of fundamental importance when COVID-19 was suspected (79.2% of the patients had a suggestive test), as well as for the outcome. The presence of about 20% of chest X-rays without any changes is consistent with what has been reported in the literature [25]. That there is such a readily performed test that predicts severity allows courses of treatment to be quickly implemented.

The most common findings on chest CTs were similar to those reported in other studies regarding the presence of ground glass opacities and septal thickening. Consolidations were slightly less common; pleural thickening and air bronchograms were much less frequently reported in this study’s cohort. By contrast, the abundance of reports of pleural effusion and atelectasis, which may be associated with other comorbidities, becomes important [26].

Although other studies have linked an increase in D-dimer with mortality as it reflects the pro-coagulant changes associated with COVID-19 [3,8,27,28], this finding was not associated with higher mortality in this study, despite a small percentage of patients who had this test available during hospitalization. This may be explained by the fact that this test was performed only when there was a clinical indication or suspicion of pulmonary thromboembolism/deep venous thrombosis (PTE/DVT) and not for the initial assessment of pro-coagulability in COVID-19.

The presence of lymphopenia was a common finding in most patients (58.4%). It was also common in other studies, despite the variation in the total leukocyte count [1, 29, 30].

A high RDW upon admission was not statistically significant as a risk factor for mortality in this cohort (31.1% vs 21.7%; RR=1.43; p=0.413). However, this finding was associated with higher mortality in the subgroup of patients aged 70 years and over (66.7% vs 33.3%; RR=2; p= 0.029). Furthermore, a high RDW during hospitalization after admission was a factor for mortality in the entire cohort (41.1% vs 10.2%; RR=4.03; p<0.001), and if all measurements are considered (admission plus elevation), having a high RDW at any time during the hospital stay was associated with higher mortality (39.7% vs 9.6%; RR=4.14; p<0.001). Some studies have associated higher RDW values with critical forms and mortality from COVID-19. [31-36] This may explain why our study found an association between RDW and mortality when age was stratified at such a high age (70 and over), where critical forms may occur more frequently, but not over the entire spectrum of the survival curve. In other words, the association was only true for higher ages. One study of 1,641 patients found a relative mortality risk of 2.73 in patients with high RDW upon admission as compared to those with normal RDW at that time. [9]

Factors that may contribute to this finding are the smaller number of patients included in the present study and the fact that there is a larger number of patients with cancer, who are known to have a better chance of having a previous change in the RDW. [37, 38] Recent studies have highlighted the prognostic value of RDW in both COVID-19 and cancer patients. In a prospective observational study, RDW was found to be a significant predictor of COVID-19 severity, with higher levels associated with severe illness and acute kidney injury (AKI). Patients with elevated RDW had increased odds of severe outcomes, emphasizing the need for routine RDW assessment in COVID-19 patients. Another study from Massachusetts General Hospital corroborated these findings, showing that an RDW greater than 14.5% at admission was linked to a significantly higher mortality risk, which remained robust even after adjusting for various demographic and clinical factors. Additionally, a further increase in RDW during hospitalization was associated with a heightened mortality risk. These insights underline the utility of RDW as a valuable biomarker for risk stratification and prognosis in both COVID-19 and cancer contexts. [39-40]

There is a simple, available, fast, and inexpensive biomarker indicative of higher mortality upon admission for the elderly as well as in disease progression for any age group: a change in the RDW at any time increases mortality in relation to normal measurements during hospitalization.

It is worth noting that these findings support considering RDW as a mortality factor even when there is a high prevalence of previous comorbidities with high clinical complexity. Even with the presence of factors for which this test may be changed prior to infection (which could explain the lack of statistical significance for said change as a risk factor for mortality at admission), this marker has a prognostic value. The relevance of including it in the risk stratification of patients hospitalized for COVID-19 is underscored, as is the fact that such an assessment ought to consider the dynamics of these changes and not only admission statistics. It also supports the use of a combination of risk factors (for example, age and RDW) in predicting the risk of negative outcomes due to COVID-19.

To the best of our knowledge, no other study has evaluated this aspect for patients of such clinical complexity for the Brazilian population. This is a single-center study with a clinical profile and previous comorbidities different from those found in other hospitals that do not provide treatment for complex cases, given that it is a university hospital known to be a reference for such cases. The race variable might interfere with hetero-identification instead of self-declaration upon admission. The number of patients with cancer may interfere with RDW values, since such an exam may already present altered values prior to SARS-CoV-2 infection in such patients. In addition, several underlying acute and chronic diseases can change the RDW, which could be a summary marker of the presence of such diseases associated with unfavorable outcomes in COVID-19.

The study concludes that elevated RDW is a significant factor associated with higher mortality in COVID-19 patients, particularly in those aged 70 years and older. A high RDW during hospitalization is linked to increased mortality across the entire cohort and having a high RDW at any point during the hospital stay correlates with higher mortality rates. Additionally, age, male sex, radiographic changes, and cardiovascular disease, which are already known severity and mortality factors, are associated with higher intrahospital mortality from COVID-19. Further studies are warranted to determine whether RDW alone can be used as a simple and available biomarker for predicting COVID-19 mortality and to explore its relationship to the pathophysiology of the disease.

## Data Availability

All data produced in the present study are available upon reasonable request to the authors

## Acknowledgement

We are deeply grateful to the patients who participated in this study and to the healthcare workers who provided unwavering care and support.

## Financial Disclosure or Funding

This research did not receive any specific grant from funding agencies in the public, commercial, or not-for-profit sectors.

## Author Contributions

Dr. Helena Duani: Conceptualization, Methodology, Supervision, Writing - original draft. Dr. Maderson Alvarez: Data curation, Formal analysis, Visualization, Writing - review & editing. Dr. Cecília Ravetti and Dr. Paula Vassallo: Investigation, Resources, Project administration. Dr. Carla Jorge <u>Machado</u>: Validation, Software, Writing - review & editing.Dr. Vandack Nobre and Milena Marcolino: Supervision, Project administration, Writing - review & editing.

## Data Availability

The data from this clinical study in the field of medicine are available upon request to the authors, subject to prior evaluation.

## Abbreviations and acronyms used in the manuscript

COVID-19: Coronavirus Disease 2019
CT: computed tomography
HC-UFMG: Hospital das Clínicas of the Universidade Federal de Minas Gerais
HR: Hazard Ratio
ICU: Intensive Care Unit
MCV: mean corpuscular volume
MV: mechanical ventilation
PNAD: National Household Sample Survey, in Portuguese: Pesquisa Nacional por Amostra de Domicílios
PTE/DVT: thromboembolism/deep venous thrombosis
RDW: Red blood cell distribution width
RR: Risk Ratio
RT-PCR: real-time polymerase chain reaction
SARS: Severe Acute Respiratory Syndrome
SD: standard deviation
SUS: Brazilian Unified Health System

